# Post-Exertional Malaise in People with Chronic Cancer-Related Fatigue

**DOI:** 10.1101/2019.12.12.19014829

**Authors:** Rosie Twomey, Samuel T. Yeung, James G. Wrightson, Guillaume Y. Millet, S. Nicole Culos-Reed

## Abstract

**Context:** Cancer-related fatigue (CRF) is a distressing and persistent sense of tiredness or exhaustion that interferes with usual functioning. Chronic CRF continues for months after curative cancer treatment is complete. Post-exertional malaise (PEM) is a worsening of symptoms after physical or mental activity, with limited investigations in people with chronic CRF.

**Objectives:** The purpose of this study was to identify and describe self-reported incidences of PEM in people with chronic CRF.

**Methods:** Participants (*n*=18) were eligible if they scored ≤34 on the Functional Assessment of Chronic Illness Therapy-Fatigue scale and had a cancer-related onset of fatigue. Participants completed a brief questionnaire to assess PEM over a 6-month time-frame (the DePaul Symptom Questionnaire – Post-Exertional Malaise; DSQ-PEM). In addition, a maximal exercise test was used to investigate self-reported symptom exacerbation (via an open-ended questionnaire) after strenuous physical exertion.

**Results:** On the DSQ-PEM, three participants met previously defined scoring criteria, which included experiencing moderate to very severe symptoms at least half of the time, worsening of fatigue after minimal effort, plus a recovery duration of >24 h. Content analysis of responses to open-ended questionnaires identified five people who experienced a delayed recovery and symptoms of PEM after maximal exercise.

**Conclusion:** A subset of people with chronic CRF (up to 33% in this sample) may experience PEM. Exercise specialists and health care professionals working with people with chronic CRF must be aware that PEM may be an issue. Symptom exacerbation after exercise should be monitored, and exercise should be tailored and adapted to limit the potential for harm.

**Key message:** This study provides preliminary evidence that a subset of people with chronic cancer-related fatigue experience post-exertional malaise.

## Introduction

Cancer-related fatigue (CRF) has been defined as a distressing, persistent sense of physical, emotional, and/or cognitive tiredness or exhaustion that is not proportional to recent activity and interferes with usual functioning [1]. The majority of people with cancer will experience CRF during active treatment, but this will resolve in the weeks after treatment. However, 20-30% of people will continue to experience CRF for months or years following treatment [2–4], Variations in estimates of prevalence may reflect variability in how CRF is defined and measured [2–4]. Many studies do not account for the time since cancer treatment or fatigue onset, meaning both transient and chronic fatigue states may be included in estimates of prevalence. Although estimates of prevalence can be high, in general, there is a natural history of improvement over time, as shown in prospective longitudinal studies where CRF is medically unexplained (i.e., not attributable to comorbidities or other factors). For example, in women treated for early-stage breast cancer, a continuous fatigue state was only present in 11% 6-months after treatment [5]. However, in a similar study in people treated for various malignancies, 22% had persistent and severe fatigue 1-year after curative cancer treatment [3]. Furthermore, higher fatigue at least 2-months after treatment predicted persistent fatigue 1-year after treatment [3]. CRF that continues for months or years has been called chronic CRF [4] or post-cancer fatigue [6]. Although there is no strict consensus on when CRF can be described as chronic, we and other research groups [6,7] define chronic CRF/post-cancer fatigue as clinically-relevant CRF that continues for ≥3 months after the completion of cancer treatment.

CRF is assessed as a patient-reported outcome, and there are psychometric tools used for both routine screening [8], and the assessment of CRF severity [9]. Evidence-based guidelines for the screening, assessment and treatment of CRF have been developed, including pan-Canadian practice guidelines for clinical use [8,10]. In addition, there are consensus-developed diagnostic criteria for CRF, as proposed for the International Statistical Classification of Diseases and Related Health Problems, 10th revision (ICD-10) in 1998 [11]. The proposed ICD-10 criteria specify that six or more symptoms must have been present every day or nearly every day during the same two-week period over the past month, and at least one symptom must be “significant fatigue.” Previous qualitative data have been interpreted as consistent with various descriptors in the ICD-10 case definition of CRF, including an excessive fatigue state, protracted in course, in relation to relatively limited physical activity [12]. Alongside items such as “perceived need to struggle to overcome inactivity” and “difficulty completing daily tasks attributed to feeling fatigued,” “post-exertional malaise (PEM) lasting several hours” is also included in the ICD-10 criteria [11]. However, PEM has not been defined in the context of CRF, and is only vaguely addressed in the accompanying diagnostic interview guide, where the related question is: “Did you find yourself feeling sick or unwell for several hours after you had done something that took some effort?” [11].

PEM is a cardinal symptom of a separate and serious long-term illness called myalgic encephalitis and/or chronic fatigue syndrome (ME/CFS) [13]. In ME/CFS, PEM is described by the Centers for Disease Control and Prevention (CDC) as a worsening of symptoms after physical or mental activity that would not have caused a problem before illness [13]. PEM in ME/CFS has also been described as a constellation of extensively disabling signs and symptoms in response to exertion [14]. Historically, there has been difficulty in defining and measuring PEM (see [15,16] for more information), but for the purposes of this study, PEM will be used to denote concepts related to post-exertional symptom exacerbation [15].

The chronic nature of CRF in some cancer survivors has led to a comparison with ME/CFS [12]. Exercise appears to exert a moderate effect on fatigue in people with CFS in the short term [17]. However, following a reanalysis of an influential study [18,19] and subsequent controversy [20,21], the CDC no longer recommends graded exercise (progressive aerobic activity) as a recommended therapy in ME/CFS [14]. However, in people living with and beyond cancer, exercise (including moderate-vigorous exercise) is considered a safe and effective intervention to counteract the adverse physical and psychological effects of cancer and its treatment [22,23], including CRF (e.g. [24]). Considering inadequate reporting of adverse reactions to exercise in previous studies of ME/CFS [17,25], and the lack of data on PEM in people with chronic CRF, caution may be warranted. PEM is inadequately documented in the literature on chronic CRF and requires further investigation considering the potential implications for exercise prescription. The purpose of this study was to identify and describe self-reported incidences of PEM in a group of people with chronic CRF. Despite the inclusion of “PEM lasting several hours” in the proposed ICD-10 criteria for CRF [6], to our knowledge, this is the first attempt to measure PEM in people with chronic CRF.

## Methods

### Participants

Participants were recruited as part of an ongoing prospective randomized controlled trial (ClinicalTrials.gov, NCT03049384 and [26]). The purpose of the RCT is to investigate the effect of a tailored versus traditional 12-week exercise intervention on fatigue severity in people with chronic CRF after cancer treatment. Participants were eligible if they were aged 18-75 years and had completed curative-intent cancer treatment (≥3 months and ≤5 years from enrolment), self-identified as feeling fatigued/tired, and self-identified that the fatigue/tiredness developed at any point during the course of the disease (e.g. during the treatment phase) and had not resolved. Furthermore, participants were eligible if they scored ≤34 [27] on the Functional Assessment of Chronic Illness Therapy Fatigue Scale (FACIT-F) [28]. The FACIT-F is widely recommended for the assessment of CRF severity [9], has a cut-off point (≤34) that correctly identifies over 90% of ‘ICD-10 positive’ cases, and has been recommended for the diagnosis of CRF [27]. Based on these eligibility criteria, all participants in the larger study were considered to have chronic CRF. Participants were excluded if they had a contraindication to experimental procedures and/or exercise. Additional inclusion and exclusion criteria have been reported previously, and include exclusion for sleep apnea or anemia [26]. A medical history was taken to account for conditions that may explain/contribute to fatigue. No participants in the present study had a diagnosis of hypothyroidism or were experiencing a major depressive episode. Written informed consent for all study procedures was obtained. This study was approved by the Health Research Ethics Board of Alberta Cancer Committee (HREBA.CC-16-10-10) and was performed according to the Declaration of Helsinki.

### Laboratory Visit

Full details of the wider project have been reported elsewhere [26]. The data for this sub-study were collected during and after an initial visit to the laboratory, previously described as Lab Visit #1 [26]. Participants completed a Physical Activity Readiness Questionnaire for Everyone (PAR-Q+) and were also screened for arrhythmia and hypertension, determined during resting electrocardiography and blood pressure measurements, respectively. Continuation with the study was conditional on the screening process, and physician approval was sought at this stage where required. Otherwise, participants were cleared for participation by a Certified Exercise Physiologist (Canadian Society for Exercise Physiology). Details of the maximal exercise test protocol have been reported in detail elsewhere [26]. Data collected during the test and used for the main study (e.g. peak oxygen uptake, heart rate, rating of perceived exertion (RPE) and blood lactate) is reported for the purpose of characterizing the test and describing the sub-study participants. Borg’s RPE scale (6-20) [29] was administered according to published instructions [30].

### Measurement of Post-Exertional Malaise

Before the exercise test, participants completed a brief questionnaire to assess PEM over a 6-month time-frame (the DePaul Symptom Questionnaire – Post-Exertional Malaise; DSQ-PEM) [31,32]. The first five items (Table 2) were recommended by the National Institutes of Health/Centers for Disease Control and Prevention Common Data Elements (CDE) PEM working group [33] and five supplementary items (including symptom duration, see Table 2) were designed to operationalize the CDE recommendations [23] further. To gain a comprehensive understanding of the participant experience, an open-ended questionnaire was provided (hard copy or via email, participant preference), and participants were instructed to complete this 96 h after the exercise test. This accounted for the potential for prolonged adverse responses as reported in people with ME/CFS [34–36], while also limiting the requirement for recall. The open-ended questionnaire was designed based on a review of the literature on PEM in ME/CFS (e.g. [12,15,34,37]) and is available online (https://osf.io/nygq5/).

**Table 1.**
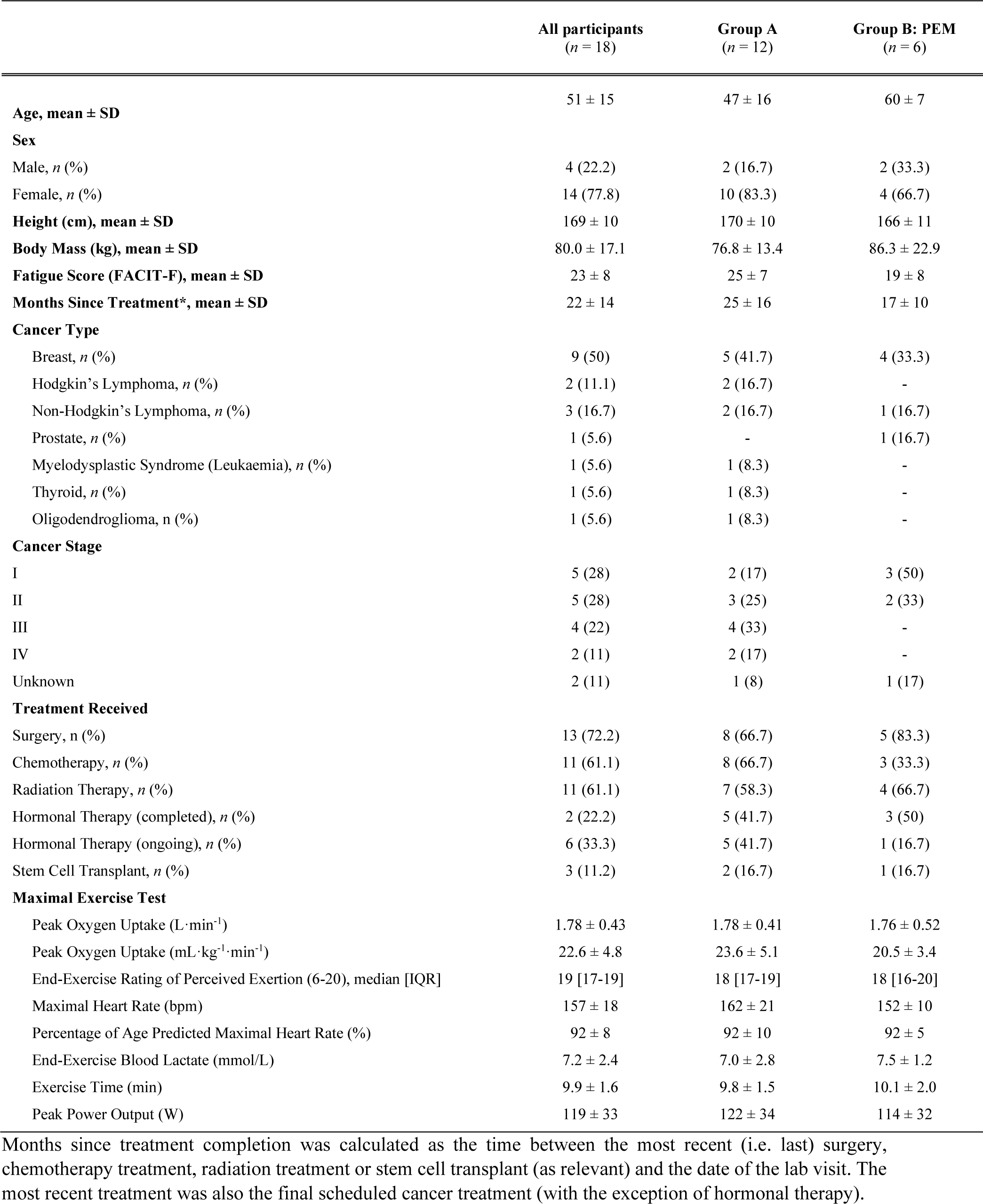
Participant Characteristics and the Maximal Exercise Test. Participant characteristics by group. Group A, participants not meeting objective criteria for the DSQ-PEM and not describing PEM in the open-ended questionnaire; Group B, participants meeting objective criteria for the DSQ-PEM and/or describing PEM in the open-ended questionnaire.

|  | All participants<br>(n = 18) | Group A<br>(n = 12) | Group B: PEM<br>(n = 6) |
| --- | --- | --- | --- |
| **Age, mean ± SD** | 51 ± 15 | 47 ± 16 | 60 ± 7 |
| **Sex** |  |  |  |
| Male, n (%) | 4 (22.2) | 2 (16.7) | 2 (33.3) |
| Female, n (%) | 14 (77.8) | 10 (83.3) | 4 (66.7) |
| **Height (cm), mean ± SD** | 169 ± 10 | 170 ± 10 | 166 ± 11 |
| **Body Mass (kg), mean ± SD** | 80.0 ± 17.1 | 76.8 ± 13.4 | 86.3 ± 22.9 |
| **Fatigue Score (FACIT-F), mean ± SD** | 23 ± 8 | 25 ± 7 | 19 ± 8 |
| **Months Since Treatment\*, mean ± SD** | 22 ± 14 | 25 ± 16 | 17 ± 10 |
| **Cancer Type** |  |  |  |
| Breast, n (%) | 9 (50) | 5 (41.7) | 4 (33.3) |
| Hodgkin's Lymphoma, n (%) | 2 (11.1) | 2 (16.7) | - |
| Non-Hodgkin's Lymphoma, n (%) | 3 (16.7) | 2 (16.7) | 1 (16.7) |
| Prostate, n (%) | 1 (5.6) | - | 1 (16.7) |
| Myelodysplastic Syndrome (Leukaemia), n (%) | 1 (5.6) | 1 (8.3) | - |
| Thyroid, n (%) | 1 (5.6) | 1 (8.3) | - |
| Oligodendroglioma, n (%) | 1 (5.6) | 1 (8.3) | - |
| **Cancer Stage** |  |  |  |
| I | 5 (28) | 2 (17) | 3 (50) |
| II | 5 (28) | 3 (25) | 2 (33) |
| III | 4 (22) | 4 (33) | - |
| IV | 2 (11) | 2 (17) | - |
| Unknown | 2 (11) | 1 (8) | 1 (17) |
| **Treatment Received** |  |  |  |
| Surgery, n (%) | 13 (72.2) | 8 (66.7) | 5 (83.3) |
| Chemotherapy, n (%) | 11 (61.1) | 8 (66.7) | 3 (33.3) |
| Radiation Therapy, n (%) | 11 (61.1) | 7 (58.3) | 4 (66.7) |
| Hormonal Therapy (completed), n (%) | 2 (22.2) | 5 (41.7) | 3 (50) |
| Hormonal Therapy (ongoing), n (%) | 6 (33.3) | 5 (41.7) | 1 (16.7) |
| Stem Cell Transplant, n (%) | 3 (11.2) | 2 (16.7) | 1 (16.7) |
| **Maximal Exercise Test** |  |  |  |
| Peak Oxygen Uptake (L·min<sup>-1</sup>) | 1.78 ± 0.43 | 1.78 ± 0.41 | 1.76 ± 0.52 |
| Peak Oxygen Uptake (mL·kg<sup>-1</sup>·min<sup>-1</sup>) | 22.6 ± 4.8 | 23.6 ± 5.1 | 20.5 ± 3.4 |
| End-Exercise Rating of Perceived Exertion (6-20), median [IQR] | 19 [17-19] | 18 [17-19] | 18 [16-20] |
| Maximal Heart Rate (bpm) | 157 ± 18 | 162 ± 21 | 152 ± 10 |
| Percentage of Age Predicted Maximal Heart Rate (%) | 92 ± 8 | 92 ± 10 | 92 ± 5 |
| End-Exercise Blood Lactate (mmol/L) | 7.2 ± 2.4 | 7.0 ± 2.8 | 7.5 ± 1.2 |
| Exercise Time (min) | 9.9 ± 1.6 | 9.8 ± 1.5 | 10.1 ± 2.0 |
| Peak Power Output (W) | 119 ± 33 | 122 ± 34 | 114 ± 32 |

Months since treatment completion was calculated as the time between the most recent (i.e. last) surgery, chemotherapy treatment, radiation treatment or stem cell transplant (as relevant) and the date of the lab visit. The most recent treatment was also the final scheduled cancer treatment (with the exception of hormonal therapy).

**Table 2.** DSQ-PEM responses.

| DSQ-PEM Item | Response | n (%) |
| --- | --- | --- |
| 1. Dead, heavy feeling after starting to exercise | Frequency $\geq 2$ and Severity $\geq 2$* | 5 (31) |
| 2. Next day soreness or fatigue after non-strenuous, everyday activities | Frequency $\geq 2$ and Severity $\geq 2$* | 7 (39) |
| 3. Mentally tired after the slightest effort | Frequency $\geq 2$ and Severity $\geq 2$* | 6 (33) |
| 4. Minimum exercise makes you physically tired | Frequency $\geq 2$ and Severity $\geq 2$* | 7 (39) |
| 5. Physically drained or sick after mild activity | Frequency $\geq 2$ and Severity $\geq 2$* | 5 (31) |
| Frequency $\geq 2$ and Severity $\geq 2$ for *at least 1* of the above items | | 9 (50) |
| Frequency $\geq 2$ and Severity $\geq 2$ for *all* of the above items | | 4 (22) |
| 6. If you were to become exhausted after actively participating in extracurricular activities, sports, or outings with friends, would you recover within an hour or two after the activity ended? | No | 8 (44) |
| 7. Do you experience a worsening of your fatigue/energy related illness after engaging in minimal physical effort? | Yes | 7 (39) |
| 8. Do you experience a worsening of your fatigue/energy related illness after engaging in mental effort? | Yes | 12 (67) |
| 9. If you feel worse after activities, how long does this last? | >24 h | 4 (22) |
| 10. If you do not exercise, is it because exercise makes your symptoms worse? | Yes | 5 (31) |

\*Frequency = “how often you have had this symptom” and a score of $\geq 2$ (2-4) ranges from “about half the time” to “all of the time.” Severity = “how much has this symptom bothered you,” and a score $\geq 2$ (2-4) ranges from “moderate” to “very severe.”

### Data Analysis

For the DSQ-PEM, items 1-5 were scored according to Cotler *et al*. [31], i.e., a frequency score of ≥2 (i.e., “about half the time” or more frequently) in combination with a severity score of ≥2 (i.e. “moderate” severity or more severe) was considered relevant. Items 6-10 are considered supplementary but were also scored according to Cotler *et al*. [31].

The open-ended questionnaires were analyzed with qualitative content analysis [38]. The written responses were de-identified, and two authors independently read all responses several times in order to become familiarized with the data. Responses were assigned codes according to a classification scheme that was developed using (i) the questions on the open-ended questionnaire/knowledge about PEM (e.g. experience of worsening of fatigue) and (ii) recurrent responses that arose from the data (e.g. the need to nap). Responses were then coded independently, and an agreement was reached on all items. Coded responses were then counted and summarized descriptively.

## Results

Of 18 participants (14 female), two did not complete the DSQ-PEM (missing data), and another completed the exercise test but did not complete the open-ended questionnaire (due to food poisoning). Participant characteristics and parameters related to the maximal exercise test are presented in Table 1.

### DSQ-PEM

In this sample of people with chronic CRF, 50% of respondents indicated that they experienced at least one of the first 5 items (see Table 2) at least half time at a moderate or greater severity. A sub-set of participants (4/16) indicated that they feel worse after activities for >24 h, and 5/16 indicated that they do not exercise because exercise makes their symptoms worse. In order to apply previously defined criteria to help identify participants for whom PEM could be an issue, all scoring steps recommended by Cotler *et al* [31] were used to identify people who scored ≥2 for frequency and severity for any item from 1-5, plus answered “Yes” to item 7 or 8, plus gave a response of >14 h for item 9. Three people in this sample (19%) met these criteria (and all three answered ≥2 for frequency and severity on all items from 1-5, answered yes for both items 7 and 8, and gave a response of >24 h for item 9).

### Open-Ended Questionnaire

There were 12 codes used for the content analysis: (1) worsening of fatigue; (2) duration/recovery time; (3) change in routine; (4) reduced daily activities; (5) muscle soreness; (6) weakness (7) joint pain; (8) mood; (9) memory/concentration; (10) need to nap or lie down; (11) flu-like symptoms; (12) positive comments on exercise/the test. A narrative synthesis of codes with illustrative quotes is provided below. Overall, we identified five participants (29% of this sample) who described PEM on the open-ended questionnaire. Specifically, these participants indicated that they experienced worsening of fatigue, plus had to change routine or reduce daily activities, plus had to nap or lie down, plus experienced ≥3 other symptoms (including muscle soreness, weakness, joint pain, mood disturbance, memory/concentration or flu-like symptoms), plus had a delayed recovery of >2 days.

Of 17 participants, 13 reported a worsening of fatigue after the exercise test. Responses ranged from mild, e.g. “*Felt a little more tired the day after the lab visit”* to more severe, e.g. “*I was thoroughly exhausted.”* Of the 13 participants who experienced worsening of fatigue, seven participants had to reduce their daily activities, and six had to change their routines or plans as a result of how they felt after the exercise test. This ranged from relatively mild: “*The only change was lying down in the morning, which is unusual for me. Typically, I get through the morning and rest in some way after lunch*.” To more severe “*The next day I stayed in bed most of the day and evening. Unable to walk the dog for two days. On the third day, I cancelled a ‘coffee date’ with a friend*.” In an item that arose from the data, six people noted that they need to nap or lie down later on the same day of the test and/or in the following days, e.g. “*I lay down all 4 afternoons for a rest. I don’t usually do that*”. Interestingly, in response to a question on the need to change plans or reduce activities, one participant answered, “*No. I anticipated the fatigue*”, indicating that they expected increased fatigue (specifically, the day after the exercise test) and had planned accordingly. Also of note, is one participant who did not need to change their routine because their routine is already reduced/disrupted by the fluctuation and unpredictable nature of their fatigue: “*I don’t usually plan anything because I don’t know how I’m going to feel especially next day, [I] only plan day to da*y.”

It was difficult to obtain clear information on how long it took for participants fatigue to return to ‘usual’ or ‘normal,’ partly because some people were unsure about their ‘usual’ or ‘normal,’ e.g. in the case of someone who only felt more tired later on the day of the test, they also commented: “*I don’t know what is abnormal for me, as things are variable*.” Another confounder was that three participants were unsure whether to attribute increased fatigue to the test, or to other activities that were perceived as strenuous that occurred within the data collection period, specifically, e.g. “*more activities with family*,” and “*more stairs than usual*.” Nevertheless, of the 12 people who indicated worse fatigue, two participants only experienced this on the day of the test, seven returned to typical levels within 1-3 days, e.g. “*Back to normal after the two days had passed*.” and four were unsure if they had recovered by 96 h (when the questionnaire was completed), e.g. “*Each evening, I seemed to be so tired that I couldn’t do my normal activities (research on family history). I didn’t have the energy or ability to concentrate. I don’t know if I have recovered that yet because I have very little energy left by evening.”*

Of 17 participants, 11 experienced muscle soreness after the test, localized to the legs and mainly attributed to cycling in all but one participant, e.g. “*The pain was just mild muscle pain that a person feels after working muscles more than usual*.” Four people experienced joint pain, and five people referred to weakness. In one person, this was prolonged: “*The weakness was extreme and lasted in its worst state for days 3 and 4*.” Seven people experienced changes in mood, and seven people had worse memory/concentration, e.g. “*First day, I had significant problems with my short-term memory and concentration.”* It is also noteworthy that one of the participants who did not report worsening of memory and concentration compared to ‘normal’ stated that they “*never feel normal*,” referring to ongoing issues with memory and concentration since cancer treatment. Five people experienced flu-like symptoms (e.g. “c*hills, cold, body wet, and very sore*” and “*I felt generalized aching throughout my body, like the flu*.”). Finally, in the additional comments, four participants self-reported feeling better after exercise in general, e.g. “*Usually feel better after exercise*” or after the test, e.g. “*I think in the afternoon I felt invigorated and then the next two days quite a bit more tired*,” and two people commented on the test itself, e.g. *“I felt very supported and safe throughout the test.”*

### Overlap Between the DSQ-PEM and Open-Ended Questionnaire

In this sample, up to 33% of participants were identified as potentially experiencing PEM (i.e. met the previously defined criteria for the DSQ-PEM and/or described PEM in the open-ended questionnaire, Table 1). Two of the five participants who described PEM in their responses to the open-ended questionnaire also met the DSQ-PEM criteria. Of the remaining three participants, one did not complete the DSQ-PEM (missing data); a second met all DSQ-PEM criteria except they answered “No” to items 7 and 8 which refer to minimal effort; and the third indicated “next day soreness or fatigue after non-strenuous, everyday activities” of a moderate severity about half the time and “minimum exercise makes you physically tired” at 2 (“severe” but only “a little of the time”), plus “yes” to items 7 and 10 (experience a worsening of fatigue after engaging in minimal physical effort, and do not exercise because it makes their symptoms worse). Although these latter two participants did not meet all scoring criteria for the DSQ-PEM, PEM does seem to be an issue above a certain intensity (i.e., above minimal), possibly explaining why the open-ended questionnaire identified these participants when the DSQ-PEM did not. Finally, the DSQ-PEM identified one participant who was not identified from the open-ended questionnaire. This participant did report increased fatigue for 2-3 days after the exercise test but also noted, “*I like exercise. I like feeling strong. Like I am working toward health”*. However, considering our experience of their later participation in the RCT, it is our clinical judgement that PEM was an issue for them (which also prevented returning to work).

## Discussion

The purpose of this study was to investigate self-reported incidences of PEM in people with chronic CRF. Our results provide preliminary evidence that a subset (up to 33% in this sample, a possible 6/18 participants) of people with chronic CRF experience symptoms of PEM. These findings must be replicated/extended in larger and more diverse samples of people with chronic CRF. Nevertheless, this finding has potential implications for exercise professionals working with people experiencing chronic CRF. We recommend that CRF is monitored throughout an exercise program (including asking people about their symptoms between individual exercise sessions, and after increases in, e.g. exercise intensity - with an awareness that symptom onset may vary and may be delayed) and that trials involving people with chronic CRF report this patient-level data to facilitate the understanding of PEM in this context, and therefore help mitigate the potential for harm.

Content analysis of open-ended questionnaires delivered 96 h after a maximal exercise test provided evidence of a constellation of symptoms, change in routine/plans, and duration of recovery in people experiencing chronic CRF. Although many participants experienced a worsening of fatigue, based on each participant’s profile of responses, and with particular consideration of the severity of symptoms and duration of their recovery, we identified five people whose descriptions were consistent with PEM. Despite qualitative comparisons between chronic CRF and ME/CFS [12,39] and the inclusion of “PEM lasting several hours” in the proposed ICD-10 criteria for CRF [11], to our knowledge, this is the first post-exercise symptom characterization reported in people experiencing chronic CRF. A number of previous studies have used maximal exercise tests to investigate the effects of physical exertion on ME/CFS symptoms (e.g. [34–36]). For example, prolonged adverse responses to maximal exercise tests are common in women with CFS (96% took >2 days to recover) and largely absent in a sedentary control group (where 87% indicated full recovery within 24 h and 100% by 2 days) [34]. Although a maximal exercise test may be more strenuous than a typical exercise session performed in line with the updated international exercise guidelines for cancer survivors [23], the DSQ-PEM asks questions relating to minimal physical effort and non-strenuous, everyday activities. Thus, we do not know what level of exertion provokes PEM in people with chronic CRF. In people with ME/CFS, low-intensity walking [40], moderate-intensity continuous training and high-intensity interval training exacerbate fatigue [41].

The current study was conceived due to the need to reduce exercise intensity and carefully monitor symptom exacerbation in some participants with chronic CRF in an ongoing exercise trial [26]. In that trial, three participants described overwhelming exhaustion between exercise sessions; therefore, little to no progression in exercise intensity or duration was made. These anecdotal descriptions are in line with recent qualitative data from Penner *et al*. [42], where fatigue was described not only as a chronic experience of imbalance but as involving “energy crashes” that forced participants to “radically reshape their day-to-day lives.” In ME/CFS, one approach that is recommended as part of a tailored and multicomponent intervention is activity pacing. Activity pacing is a self-management strategy that encourages the individual to be as active as possible but to rest or switch activities in response to internal cues, in order to avoid marked exacerbations of symptoms (for a review, see [43]). One study added activity pacing as part of a multicomponent intervention (including graded exercise therapy) in chronic CRF, and 32% of participants had a clinically-significant improvement in fatigue [6]. Activity pacing may be beneficial for individuals with chronic CRF, not only in relation to exercise but for the management of day-to-day activities.

In this study, 33% of participants indicated that they became “mentally tired after the slightest effort” (DSQ-PEM, Table 2). This is in line with previous findings, and cognitive symptoms reported in people with chronic CRF may be indistinguishable from those reported in people with ME/CFS [39,44]. Furthermore, after the exercise test, some symptom exacerbation was cognitive in nature, i.e. impaired concentration or short-term memory. This is also in line with previous research in ME/CFS (e.g.[35,45]). Considering that no only physical, but also cognitive exertion can result in PEM in people with ME/CFS [16,35], it is possible that this could also be the case in a subset of people with chronic CRF.

Although commentary about individuals in this study is anecdotal, it is important to communicate about individual profiles to minimize the potential for harm. Clearly, nuance is required in the interpretation of these two PEM measurements. Our goal is increased awareness of PEM as a potential issue within the wider context of “exercise is medicine” in oncology [46]. If exercise is medicine in chronic CRF, side-effects at a given exercise dose must be carefully detailed. This will enable exercise professionals to provide the best exercise prescription in people with PEM, one that can be achieved within a pacing strategy that focusses on balancing rest and activity to achieve daily tasks, maintaining physical function, and avoiding symptom exacerbation. The new exercise guidelines for cancer survivors [23] acknowledge that some proportion of cancer survivors may not be able to tolerate the recommendations (particularly because dose reductions may have been poorly reported in previous trials). The results of this study reinforce the need to monitor tolerance and symptom exacerbation, especially in those with chronic CRF. Without appropriate safeguards (e.g. monitoring of fatigue throughout the exercise intervention), it is not known if people with PEM continue with little symptom exacerbation (e.g. due to low effort, or individualization and regular communication with an exercise professional), continue despite symptom exacerbation (and experience “energy crashes” [42] that may significantly impact daily activities), or drop out. It has been recently highlighted that the reporting of exercise interventions and specifically the “precise reporting of exercise treatment adherence (tolerability)” [47] must be improved. We add that these reports must include the worsening of symptoms, including fatigue. However, it should be emphasized that, for the majority of people with chronic CRF, exercise is likely to be beneficial for multiple and interacting components of physical and mental health. With support, most participants can reach physical activity guidelines (e.g. 30 minutes of aerobic activity at RPE 12, three times per week [23]) with no adverse consequences. Higher intensities/longer durations can be appropriate for people with chronic CRF who do not experience any adverse responses, particularly if the goal is to maximize physiological adaptations.

### Study Limitations and Future Directions

In Table 1, descriptive differences are provided for participants not meeting objective criteria for the DSQ-PEM and not describing PEM in the open-ended questionnaire (Group A, *n* = 12); and participants meeting objective criteria for the DSQ-PEM and/or describing PEM in the open-ended questionnaire (Group B, *n* = 6). Although of interest, the small sample size in Group B precludes in-depth interpretation. Although of interest, the small sample size in Group B precludes in-depth interpretation. Future research should follow-up on the potential effects of age, fatigue severity, time since treatment, and the type or combination of treatment received on PEM. We note that for one individual categorized to Group B (Table 1), the recall period of the DSQ-PEM (6 months) overlapped with the completion of adjuvant therapy (4 months prior to data collection) and this may have influenced their responses. In line with studies in ME/CFS [35,41,48], future studies on PEM in people with chronic CRF should record symptoms at several time points (e.g. 24, 48 and 72 h post-exercise) to better capture the symptom trajectory. This study did not include a control group or data for the responses of healthy/sedentary adults because this was outside the scope of our research question. However, in a previous study, 100% of controls recovered within 2 days of a maximal exercise test, 87% experienced an increased sense of wellbeing relative to the test, and none experienced issues such as weakness, impaired memory/concentration or mood disturbance [34]. The DSQ-PEM is not validated for use in people with CRF. However, it has been used in people with multiple sclerosis and post-polio syndrome (other clinical conditions where chronic fatigue can be an issue) [31]. As stated previously, the DSQ-PEM was not used as a method to support a diagnosis of ME/CFS study, especially considering the diagnostic complexity of ME/CFS [49]). Here, it is used as a tool to help identify people experiencing symptoms of PEM, and any further speculation on whether participants would meet any given diagnostic criteria for ME/CFS if assessed by an expert clinician is out of our scope of practice. Because we were unable to obtain a copy of the open-ended questionnaire used previously [34], an open-ended questionnaire was designed (https://osf.io/349ky/) based on a review of the literature on PEM in ME/CFS. Another limitation is that maximal incremental exercise is not a typical exercise intensity prescribed within an intervention, and provides no information about the intensity of physical effort (below maximal) that could result in PEM. Furthermore, although the FACIT-F has been recommended to support a diagnosis of CRF [50], there is no consensus on what “diagnosis” means in this context. The proposed ICD-10 criteria were not accepted, and few studies have implemented them [51], and current guidelines have differing recommendations for screening and assessment [8,52], and we recognize a lack of consensus for the identification of someone with clinically-relevant CRF. Further, we acknowledge that the inclusion/exclusion criteria for this study were not equivalent to the extensive investigations/assessments that inform a diagnosis of ME/CFS

In this preliminary study, up to 33% of people appeared to experience PEM, but all participants self-selected themselves into an RCT that explicitly involved a 12-week exercise intervention. It is reasonable to suggest that the sample in the RCT, and therefore this sub-study, may not represent the prevalence of PEM in people with chronic CRF, because people who experience PEM may be less likely to enrol in an exercise trial. Future studies should estimate the prevalence of PEM in people with chronic CRF. In addition, future studies should include semi-structured qualitative interviews to elucidate the cancer-specific symptoms and impact of PEM. This preliminary study included people with chronic CRF, identified using a cancer-related symptom onset, and presence of CRF at least 3 months and within 5 years (22 ± 14 months) after curative-intent cancer treatment. Because CRF is more prevalent during cancer treatment [53], future work must address the potential of PEM after exercise in interventions delivered during treatment. One study pre-emptively reduced the exercise prescription to encourage attendance when treatment side effects (including fatigue) were anticipated to peak during a chemotherapy cycle [54], but to our knowledge, there has been no post-exertional symptom characterization during cancer treatment.

## Conclusion

This study provides preliminary evidence that a subset of people with chronic CRF experience PEM. We recommend that exercise professionals not only monitor fatigue throughout an exercise program (i.e. asking participants how they felt in between individual sessions - with an awareness that symptom onset may vary and may be delayed) but also report this data if the exercise program is part of a clinical trial. In conclusion, the field of exercise oncology must be aware that adverse responses to exercise may be an issue for a subset of people with chronic CRF, and exercise interventions should be designed and adapted to mitigate symptom exacerbation and limit the potential for harm.

## Disclosure/Conflict of Interest

This research was supported by the Canadian Cancer Society (grant #704208-1). The authors have no conflicts of interest to disclose.

## Data Availability

There is no statistical reporting in this manuscript (therefore, no statistical code is provided). Data from the DSQ-PEM and qualitative data is summarized narratively within the manuscript.

## Notes

### Competing Interest Statement

The authors have declared no competing interest.

### Clinical Protocols

https://osf.io/nygq5/

### Funding Statement

This research was funded by the Canadian Cancer Society (grant #704208-1).

